# Mortality Risk and Temporal Patterns of Atrial Fibrillation in the Nationwide Registry

**DOI:** 10.1101/2021.01.30.21250715

**Authors:** Sirin Apiyasawat, Sakaorat Kornbongkotmas, Ply Chichareon, Rungroj Krittayaphong, for the COOL-AF Investigators

## Abstract

**Aims:** Persistent and permanent atrial fibrillation (AF) often occurs in the presence of multiple comorbidities and is linked to adverse clinical outcomes. It is unclear whether the sustained pattern of AF itself is prognostic or if it is confounded by underlying comorbidities. Here, we tested the association between the temporal patterns of AF and the risks of ischemic stroke and all-cause mortality.

**Methods and Results:** In a prospective multicenter cohort, 3046 non-valvular AF patients were consecutively enrolled and followed for adverse outcomes of all-cause mortality and ischemic stroke. The risks of both outcomes were adjusted for underlying comorbidities, and compared between the patterns of AF. At baseline, the patients were classified as paroxysmal (N=963, 31.6%), persistent (N=604, 19.8%), and permanent AF (N=1479, 45.6%) according to the standard definition. Anticoagulants were administered in 75% of all patients and 81% of those with CHA_2_DS_2_-VAS_c_ ≥2. During a mean follow up of 26 (SD 10.5) months, all-cause mortality occurred less in paroxysmal AF (2.5 per 100 patient-years) than in persistent AF (4.4 per 100 patient-years; adjusted hazard ratio [HR] 0.66, 95% CI, 0.46-0.96; *P* = .03) and permanent AF (4.1 per 100 patient-years; adjusted HR 0.71, 95% CI, 0.52-0.98; *P* = .04). The risk of ischemic stroke was similar across all patterns of AF.

**Conclusions:** In this multicenter registry of well-anticoagulated AF patients, persistent and permanent AF was associated with higher all-cause mortality than paroxysmal AF, independent of baseline comorbidities.

**Clinical Trial Registration:** Thai Clinical Trial Registration; Study ID: TCTR20160113002

## Introduction

Atrial fibrillation (AF) is categorized into paroxysmal, persistent, and permanent according to temporal patterns. Studies have been shown that approximately one-third^1^ to one-fourth^2^ of patients with paroxysmal AF progress to persistent AF within 10 years. This ratio is even higher in the presence of structural heart disease—reportedly more than half progressing in 10 years.^3^ Those with sustained forms of AF are linked to higher comorbidities and worse clinical outcomes.^2,4-7^

Current recommendation for management of AF focuses on controlling comorbidities to reduce adverse events rather than interrupting the progression of AF.^8^ However, recent trials suggested that the temporal pattern of AF itself may be an independent predictor of clinical outcomes.^4,9^ Persistent form of AF can lead to both atrial and ventricular remodeling that could subsequently progress to AF-mediated cardiomyopathy.^10-12^ Interrupting the progression of AF at the early stage has also recently been shown to reduce stroke and cardiovascular death.^13^

Here, we aim to investigate the association between the temporal patterns of AF and adverse clinical outcomes in the COOL-AF registry (**<u>Co</u>**hort of Antithrombotic Use and **<u>O</u>**ptimal INR **<u>L</u>**evel in Patients with Non-Valvular **<u>A</u>**trial **<u>F</u>**ibrillation in Thailand), a nationwide prospective cohort of AF patients. We hypothesize that paroxysmal AF was associated with lower mortality and stroke risks than persistent or permanent AF.

## Methods

### Study Population

COOL-AF registry is a multicenter prospective cohort study of patients with non-valvular atrial fibrillation (NVAF). The study consecutively enrolled patients from 27 hospitals^14^ across all regions of Thailand. The inclusion and exclusion criteria were described previously.^14^ Briefly, adults (ie, age >18) with electrocardiography-confirmed AF were eligible for the enrolment. The exclusion criteria were (1) ischemic stroke within 3 months; (2) hematologic disorders that can increase the risk of bleeding such as thrombocytopenia (<100000/mm^3^) and myeloproliferative disorders; (3) mechanical prosthetic valve or valve repair; (4) rheumatic valve disease or severe valve disease; (5) AF associated with transient reversible cause; (6) current participation in a clinical trial; (7) life expectancy <3 years; (8) pregnancy; (9) inability to attend follow-up visits; and (10) refusal to participate in the study. The protocol was approved by the ethics committee of each participating hospital. All patients provided written informed consent.

### Data Collection

The source of data were medical records and patients or family interviews. Data collected at baseline included demographic profile, body weight and height, temporal pattern of AF, medical history, clinical examination, laboratory data, medication use, and all components CHA_2_DS_2_-VAS_c_ and HASBLED scores. Temporal patterns of AF were classified by investigators as paroxysmal (AF terminated within 7 days), persistent (AF sustained >7 days), and permanent (continuous AF > 12 months with failed attempts to restore sinus rhythm) according to the standard guideline.^15^ All patients were prospectively followed for 36 months. Those with newly diagnosed AF and those with incomplete data were excluded from this analysis.

The enrolment and all data acquisition occurred between 2014 – 2017. All data were entered in a predefined web-based form and centrally validated for completion. Monitoring visits were conducted at all participating sites to ensure compliance with the protocol and validity of the data.

### Assessment of Clinical Outcomes

Patients were prospectively followed for the primary outcomes of all-cause mortality and ischemic stroke. The secondary outcomes were cardiovascular death, major bleeding, and intracranial hemorrhage. The occurrence of these events was monitored every 6 months until the end of follow-up at 36 months. All events were validated by the adjudication committee. Ischemic stroke was defined as an episode of neurological dysfunction caused by focal cerebral, spinal, or retinal infarction.^16^ Major bleeding was defined according to the International Society of Thrombosis and Haemostasis criteria,^17^ which were fatal bleeding, bleeding in a critical area or organ(s), bleeding that results in a decrease in hemoglobin level of at least 20 g/L, and bleeding that requires a transfusion of at least two units of red cells.

### Statistical Analysis

Continuous data were reported as mean (SD); categorical data, as number (percentage). The event rates of the clinical outcomes were measured as patient-year. Differences in baseline characteristics between the patterns of AF were compared by using One-way ANOVA with post-hoc least significant difference test (LSD) for continuous variables and Chi-square test with post-hoc pairwise comparisons for categorical variables. Kaplan-Meier with log-rank test was used to estimate the survival differences between the patterns. Hazard ratios (HRs) with 95% confidence intervals (CI) of clinical outcomes were calculated by Cox proportional hazards models. Using backward step-wise selection method, the models were adjusted with age, sex, body mass index, smoking status, alcohol use, hypertension, dyslipidemia, bleeding history, congestive heart failure (CHF), diabetes mellitus, history of stroke or transient ischemic attack (Stroke/TIA), vascular disease, abnormal renal function, abnormal liver function, use of antiplatelet, and use of anticoagulant. Validation for proportional hazards assumption was performed using Schoenfeld tests for all global models and scaled Schoenfeld residuals for each covariate.^18^ There were no violations detected graphically or by Grambsch-Therneau tests (ie, all *P* values for global models and for each covariate were > .05). Statistical adjustment for multiplicity was not performed due to the exploratory nature of this trial.^19^ A *P* value of < .05 was considered significant. All analyses were conducted using IBM SPSS Statistics for Windows, Version 25.0 (Armonk, NY: IBM Corp.) and Stata Statistical Software: Release 13 (College Station, TX: StataCorp LP).

## Results

### Baseline Findings

A total of 3402 patients were enrolled in the COOL-AF registry. After excluding the patients with newly diagnosed AF (N=91) and those with incomplete data (N=275), 3046 patients (mean age 67.1, 41% female, mean CHA_2_DS_2_-VAS_c_ score 2.97) remained in the analysis. Of those, 963 patients (31.6%) were classified as paroxysmal AF, 604 patients (19.8%) as persistent AF, and 1479 patients (48.6%) as permanent AF (Table Patients with paroxysmal AF were younger (mean age 65.9 years) than those with persistent AF (mean age 66.8 years) and those with permanent AF (mean age 68.1 years). Patients with paroxysmal AF were more likely female, less likely to have history of CHF, and had lower CHA_2_DS_2_-VAS_c_ and HASBLED scores than those with persistent and permanent AF. History of Stroke/TIA was highest in permanent AF followed by persistent and paroxysmal AF, while bleeding history was essentially the same across all patterns of AF. The overall anticoagulation rate was 75% (N=2286) and increased to 81.5% in patients with CHA_2_DS_2_-VAS_c_ ≥ 2. Between the patterns of AF (Figure 1), the rate was higher in permanent AF (N=1192, 80.6%) than persistent (N=448, 74.2%, *P* vs permanent = .001) and paroxysmal AF (N=646, 67.1%, *P* vs permanent < .001). The dual anticoagulant and antiplatelet therapy was administered in 275 patients (9%); more often in paroxysmal (N=101, 10.5%) and persistent AF (N=70, 11.6%) than permanent AF (N=104, 7%; *P* vs paroxysmal and *P* vs persistent < .001).

**Figure 1.**
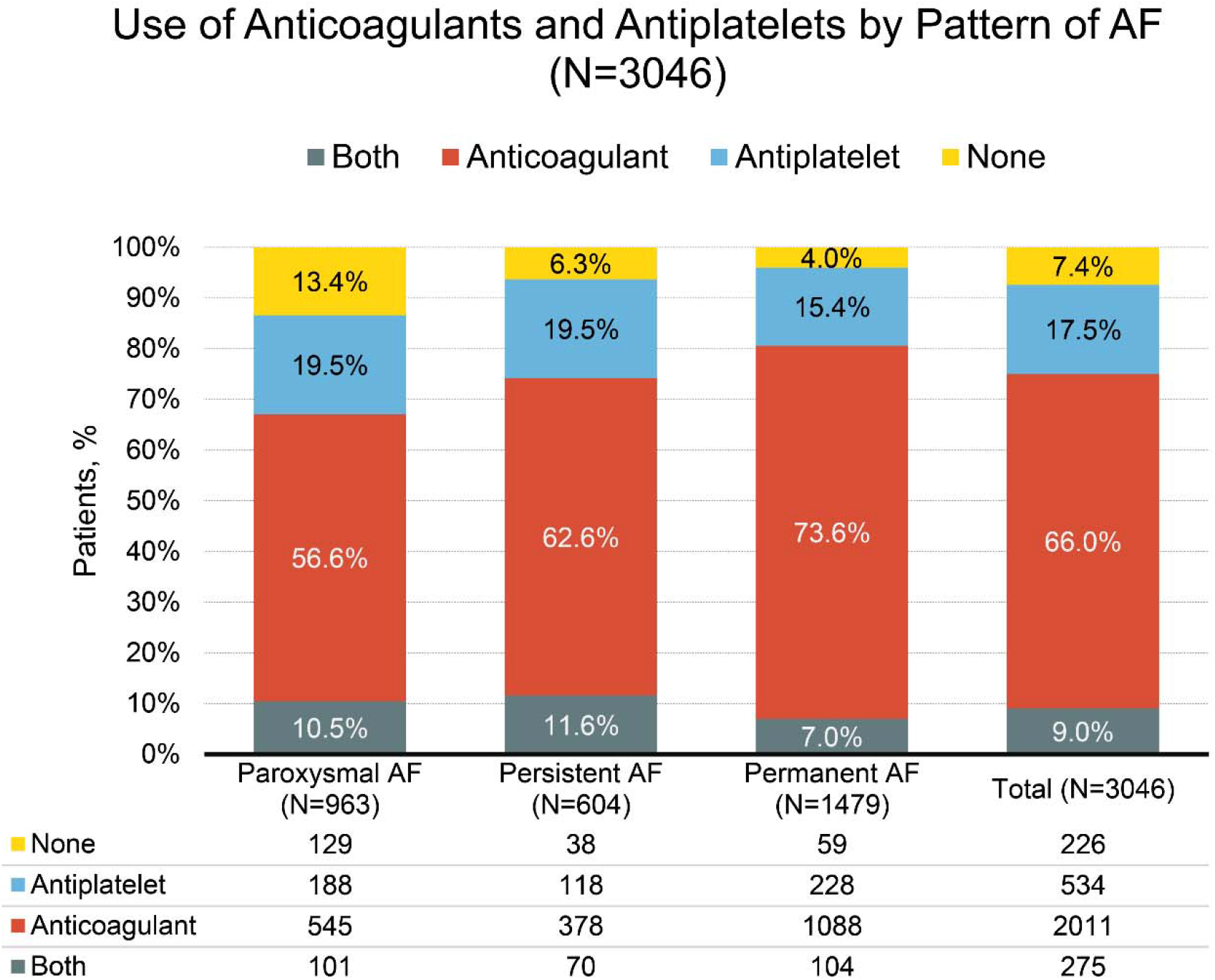
Anticoagulant and Antiplatelet Therapy by Pattern of Atrial Fibrillation. Abbreviation: AF, atrial fibrillation.

### Outcomes

During a mean follow up of 26 (SD 10.5) months, 240 patients died (7.9%; 3.6 per 100 patient-years). The causes of death (Table 2) were cardiovascular in 82 patients (34.2%), non-cardiovascular in 110 patients (45.8%), and undetermined in 48 patients (20%). Those with paroxysmal AF were less likely to die of any causes (2.5 per 100 patient-years) than those with persistent AF (4.4 per 100 patient-years; adjusted HR 0.66, 95% CI, 0.46-0.96; *P* = .03) and those with permanent AF (4.1 per 100 patient-years; adjusted HR 0.71, 95% CI, 0.52-0.98; *P* = .04; Figure 2 and Table 3). There were no differences in the incidence rates of cardiovascular death, non-cardiovascular death, ischemic stroke (Figure 3), and intracranial hemorrhage between the patterns of AF (Table 3). Major bleeding was fewer in patients with paroxysmal AF (1.5 per 100 patient-years) than those with persistent AF (2.6 per 100 patient-years; adjusted HR 0.59, 95% CI, 0.67-0.97; *P* = .04; Table 3). Comparing between persistent and permanent AF, the incidences of all-cause mortality, cardiovascular death, ischemic stroke, and intracranial hemorrhage were not statistically different.

**Table 1.** Baseline Characteristics by Patterns of Atrial Fibrillation.

|  | Px | Ps | Pm | Pair-wise *P* value |  |  |
| --- | --- | --- | --- | --- | --- | --- |
|  | (N=963) | (N=604) | (N=1479) | Px vs. Ps | Px vs. Pm | Ps vs. Pm |
| **Demographics** |  |  |  |  |  |  |
| Age, mean (SD), y | 65.9 (11.3) | 66.8 (11.1) | 68.1 (11.0) | .11 | <.001 | .02 |
| Female, No. (%) | 461 (47.9%) | 221 (36.6%) | 578 (39.1%) | <.001 | <.001 | .86 |
| BMI, mean (SD),<br>m<sup>2</sup>/kg | 25.2 (4.7) | 27.0 (28.4) | 25.1 (4.8) | .01 | .873 | .004 |
| **Medical History** |  |  |  |  |  |  |
| Current Smoker,<br>No. (%) | 20 (2.1%) | 20 (3.3%) | 58 (3.9) | .13 | .01 | .51 |
| Alcohol Use, No.<br>(%) | 31 (3.2%) | 26 (4.3%) | 71 (4.8%) | .26 | .06 | .63 |
| Hypertension, No.<br>(%) | 642 (66.7%) | 409 (67.7%) | 1032<br>(69.8%) | .67 | .11 | .36 |
| Diabetes, No. (%) | 242 (25.1%) | 147 (24.3%) | 368 (24.9%) | .72 | .89 | .79 |
| CAD, No. (%) | 162 (16.8%) | 116 (19.2%) | 202 (13.7%) | .23 | .03 | .001 |
| Stroke/TIA, No. (%) | 141 (14.6%) | 94 (15.6%) | 284 (19.2%) | .62 | .004 | .051 |
| Vascular Disease,<br>No. (%) | 168 (17.4%) | 125 (20.7%) | 215 (14.5%) | .11 | .053 | .001 |
| CHF, No. (%) | 154 (15.9%) | 172 (28.5%) | 404 (27.3%) | <.001 | <.001 | .59 |
| Abnormal Renal<br>Function, N (%) | 30 (3.1%) | 21 (3.5%) | 44 (3.0%) | .69 | .84 | .55 |
| Abnormal Liver<br>Function, No. (%) | 22 (2.3%) | 16 (2.6%) | 33 (2.2%) | .65 | .93 | .57 |
| Dementia, No. (%) | 6 (0.6%) | 4 (0.7%) | 14 (0.9%) | .92 | .39 | .53 |
| Bleeding History,<br>No. (%) | 88 (9.1%) | 52 (8.6%) | 159 (10.8%) | .72 | .20 | .14 |
| **Medications** |  |  |  |  |  |  |
| Antiplatelet, No. (%) | 289 (30.0%) | 188 (31.1%) | 332 (22.4%) | .64 | <.001 | <.001 |
| OAC, No. (%) | 646 (67.1%) | 448 (74.2%) | 1192<br>(80.6%) | .003 | <.001 | .001 |
| DOACs, No. (%) | 89 (9.2%) | 33 (5.5%) | 61 (4.1%) | .007 | <.001 | .18 |
| Antiplatelet and<br>OAC, No. (%) | 101 (10.5%) | 70 (11.6%) | 104 (7.0%) | .885 | <.001 | <.001 |
| **Risk Scores** |  |  |  |  |  |  |
| CHA<sub>2</sub>DS<sub>2</sub>-VAS<sub>c</sub>,<br>mean (SD) | 2.82 ± 1.7 | 2.95 ± 1.6 | 3.08 ± 1.6 | .12 | <.001 | .10 |
| CHA<sub>2</sub>DS<sub>2</sub>-VAS<sub>c</sub> ≥ 2,<br>No. (%) | 732 (76.0%) | 481 (79.6%) | 1230<br>(83.2%) | .09 | <.001 | .06 |
| HASBLED, mean<br>(SD) | 1.44 ± 1.0 | 1.51 ± 1.0 | 1.61 ± 1.0 | .14 | <.001 | .04 |

Abbreviations: BMI, body mass index; CAD, coronary artery disease; CHF, congestive heart failure; DOAC, direct oral anticoagulant; OAC, oral anticoagulant; Px, paroxysmal atrial fibrillation; Ps, persistent atrial fibrillation; Pm, permanent atrial fibrillation.

**Table 2.**
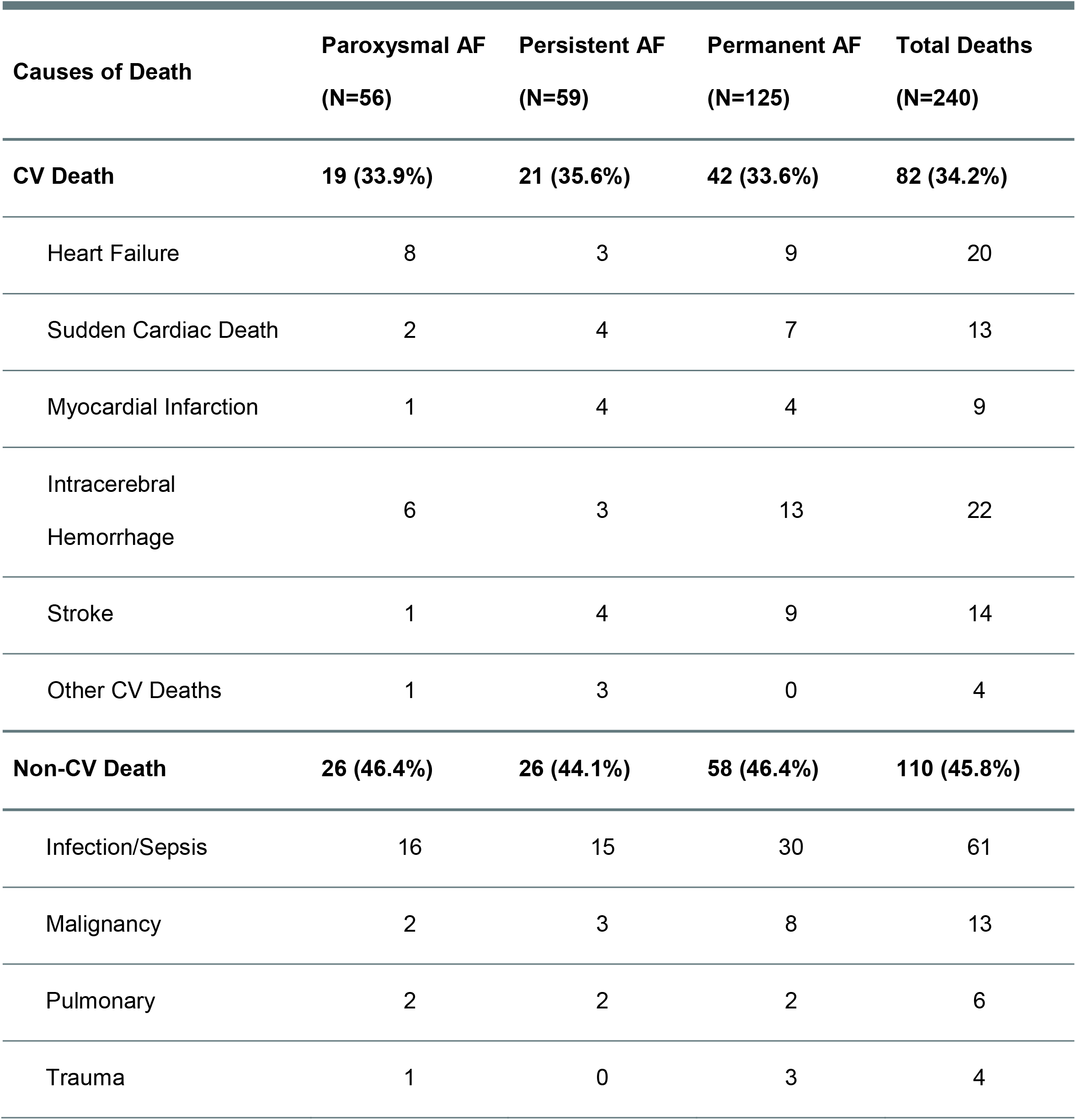
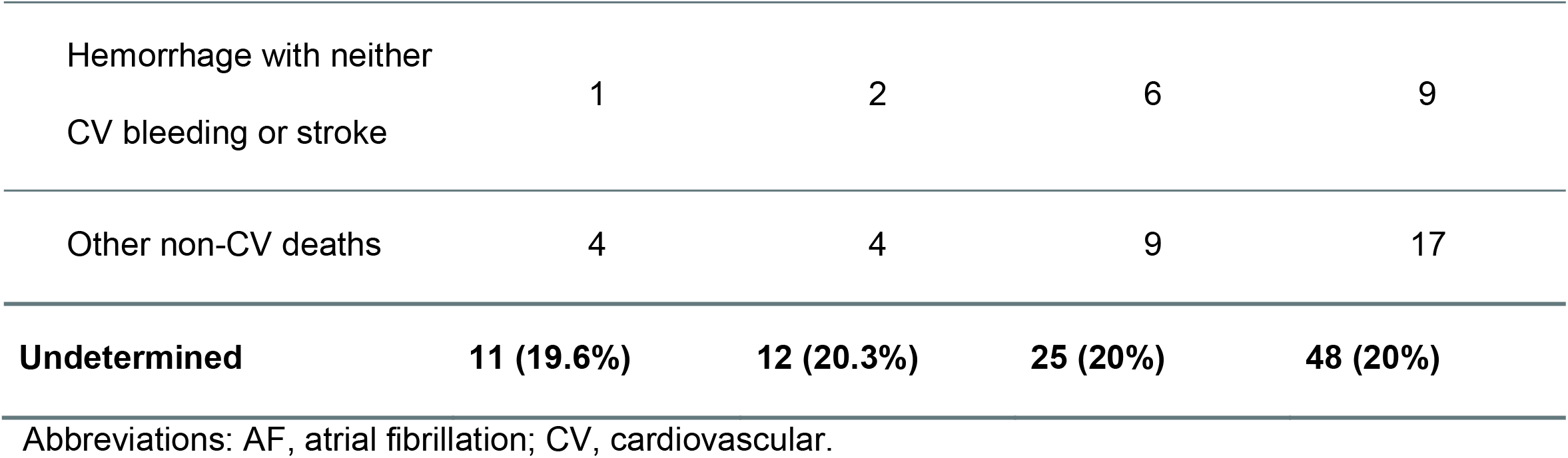
Causes of Death by Patterns of Atrial Fibrillation.

| **Causes of Death** | **Paroxysmal AF**<br>**(N=56)** | **Persistent AF**<br>**(N=59)** | **Permanent AF**<br>**(N=125)** | **Total Deaths**<br>**(N=240)** |
| --- | --- | --- | --- | --- |
| **CV Death** | **19 (33.9%)** | **21 (35.6%)** | **42 (33.6%)** | **82 (34.2%)** |
| Heart Failure | 8 | 3 | 9 | 20 |
| Sudden Cardiac Death | 2 | 4 | 7 | 13 |
| Myocardial Infarction | 1 | 4 | 4 | 9 |
| Intracerebral<br>Hemorrhage | 6 | 3 | 13 | 22 |
| Stroke | 1 | 4 | 9 | 14 |
| Other CV Deaths | 1 | 3 | 0 | 4 |
| **Non-CV Death** | **26 (46.4%)** | **26 (44.1%)** | **58 (46.4%)** | **110 (45.8%)** |
| Infection/Sepsis | 16 | 15 | 30 | 61 |
| Malignancy | 2 | 3 | 8 | 13 |
| Pulmonary | 2 | 2 | 2 | 6 |
| Trauma | 1 | 0 | 3 | 4 |
| Hemorrhage with neither<br>CV bleeding or stroke | 1 | 2 | 6 | 9 |
| Other non-CV deaths | 4 | 4 | 9 | 17 |
| **Undetermined** | **11 (19.6%)** | **12 (20.3%)** | **25 (20%)** | **48 (20%)** |

Abbreviations: AF, atrial fibrillation; CV, cardiovascular.

**Table 3.**
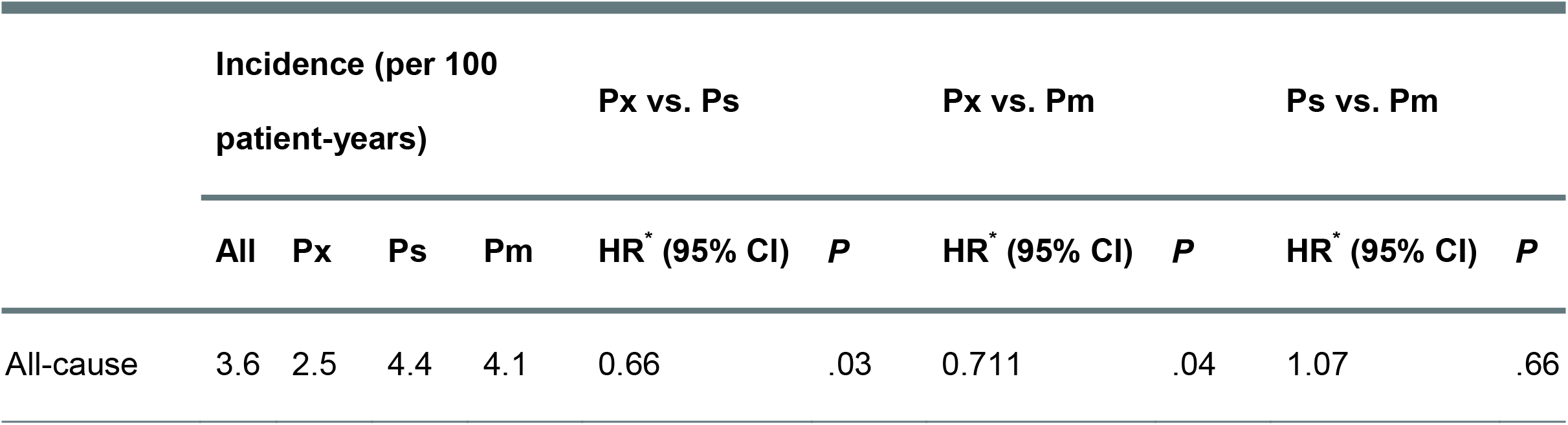
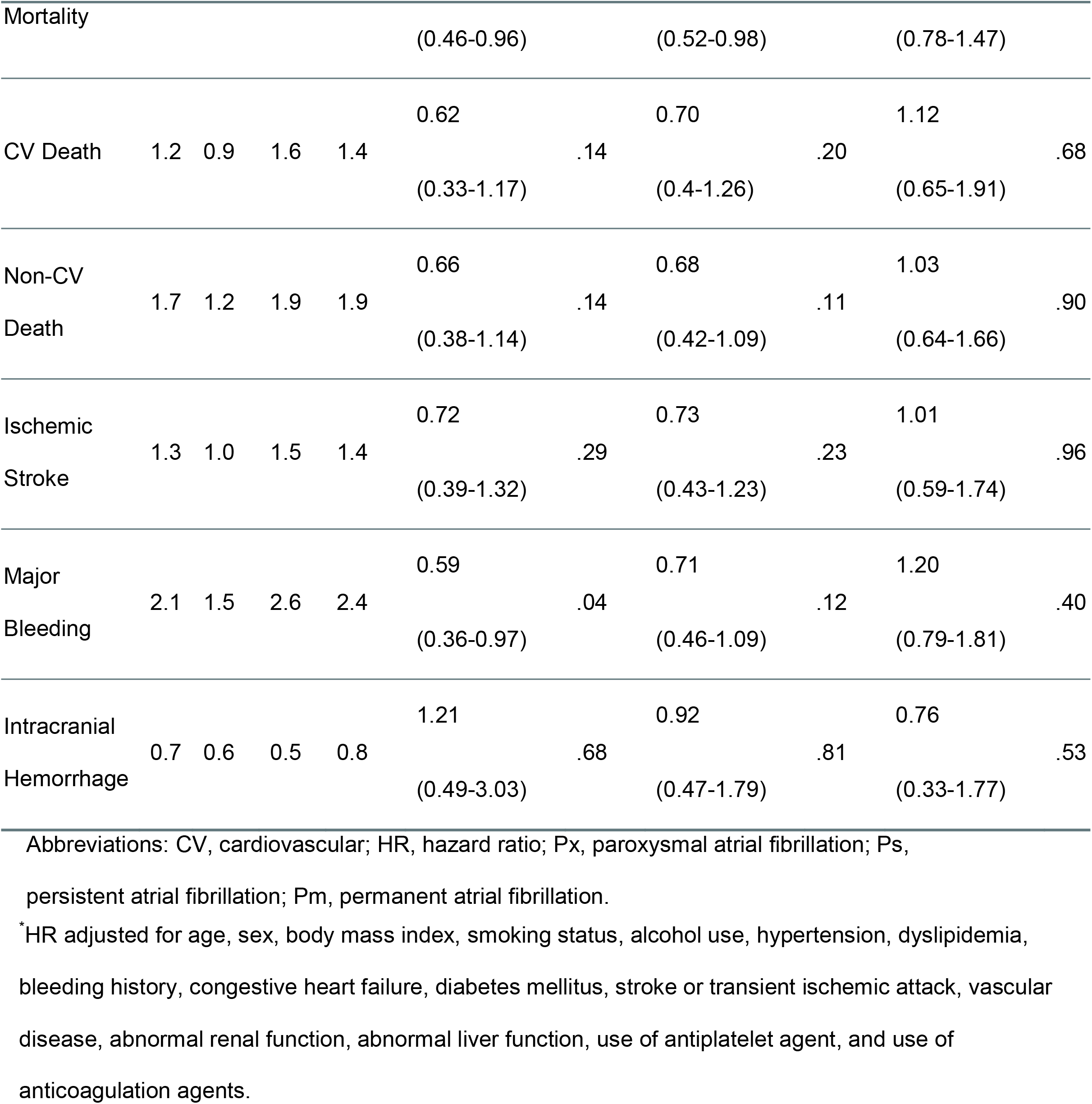
Clinical Outcomes by Pattern of Atrial Fibrillation.

|  | Incidence (per 100<br>patient-years) |  |  |  | Px vs. Ps |  | Px vs. Pm |  | Ps vs. Pm |  |
| --- | --- | --- | --- | --- | --- | --- | --- | --- | --- | --- |
|  | All | Px | Ps | Pm | HR* (95% CI) | P | HR* (95% CI) | P | HR* (95% CI) | P |
| All-cause | 3.6 | 2.5 | 4.4 | 4.1 | 0.66 | .03 | 0.711 | .04 | 1.07 | .66 |

|  | Incidence (per 100 patient-years) |  |  |  | Px vs. Ps |  | Px vs. Pm |  | Ps vs. Pm |  |
| --- | --- | --- | --- | --- | --- | --- | --- | --- | --- | --- |
|  | All | Px | Ps | Pm | HR* (95% CI) | P | HR* (95% CI) | P | HR* (95% CI) | P |
| Mortality |  |  |  |  | (0.46-0.96) |  | (0.52-0.98) |  | (0.78-1.47) |  |
| CV Death | 1.2 | 0.9 | 1.6 | 1.4 | 0.62<br>(0.33-1.17) | .14 | 0.70<br>(0.4-1.26) | .20 | 1.12<br>(0.65-1.91) | .68 |
| Non-CV Death | 1.7 | 1.2 | 1.9 | 1.9 | 0.66<br>(0.38-1.14) | .14 | 0.68<br>(0.42-1.09) | .11 | 1.03<br>(0.64-1.66) | .90 |
| Ischemic Stroke | 1.3 | 1.0 | 1.5 | 1.4 | 0.72<br>(0.39-1.32) | .29 | 0.73<br>(0.43-1.23) | .23 | 1.01<br>(0.59-1.74) | .96 |
| Major Bleeding | 2.1 | 1.5 | 2.6 | 2.4 | 0.59<br>(0.36-0.97) | .04 | 0.71<br>(0.46-1.09) | .12 | 1.20<br>(0.79-1.81) | .40 |
| Intracranial Hemorrhage | 0.7 | 0.6 | 0.5 | 0.8 | 1.21<br>(0.49-3.03) | .68 | 0.92<br>(0.47-1.79) | .81 | 0.76<br>(0.33-1.77) | .53 |

Abbreviations: CV, cardiovascular; HR, hazard ratio; Px, paroxysmal atrial fibrillation; Ps, persistent atrial fibrillation; Pm, permanent atrial fibrillation.
\*HR adjusted for age, sex, body mass index, smoking status, alcohol use, hypertension, dyslipidemia, bleeding history, congestive heart failure, diabetes mellitus, stroke or transient ischemic attack, vascular disease, abnormal renal function, abnormal liver function, use of antiplatelet agent, and use of anticoagulation agents.

**Figure 2.**
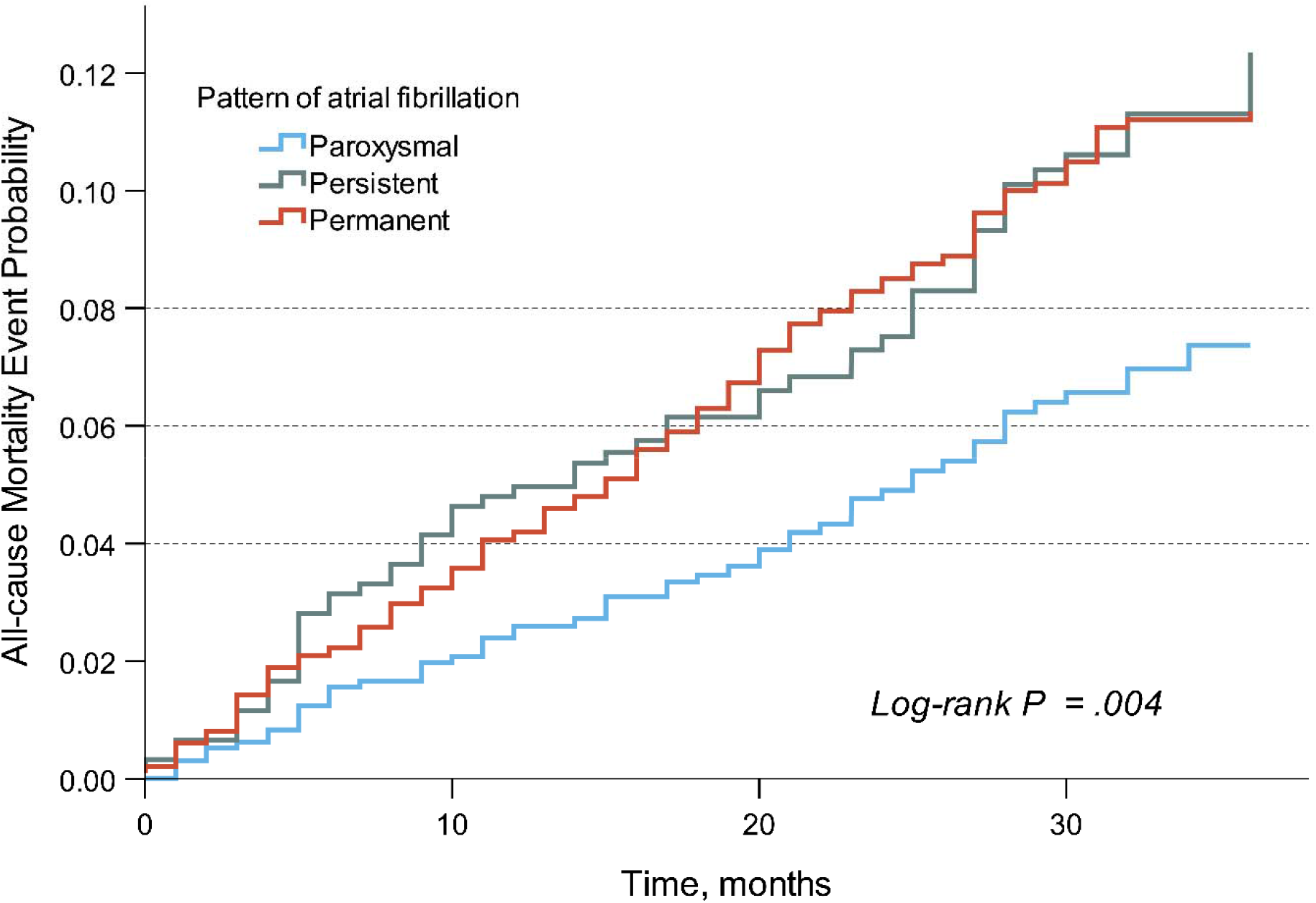
Kaplan-Meier Cumulative Probability Curve of All-cause Mortality by Pattern of Atrial Fibrillation.

**Figure 3.**
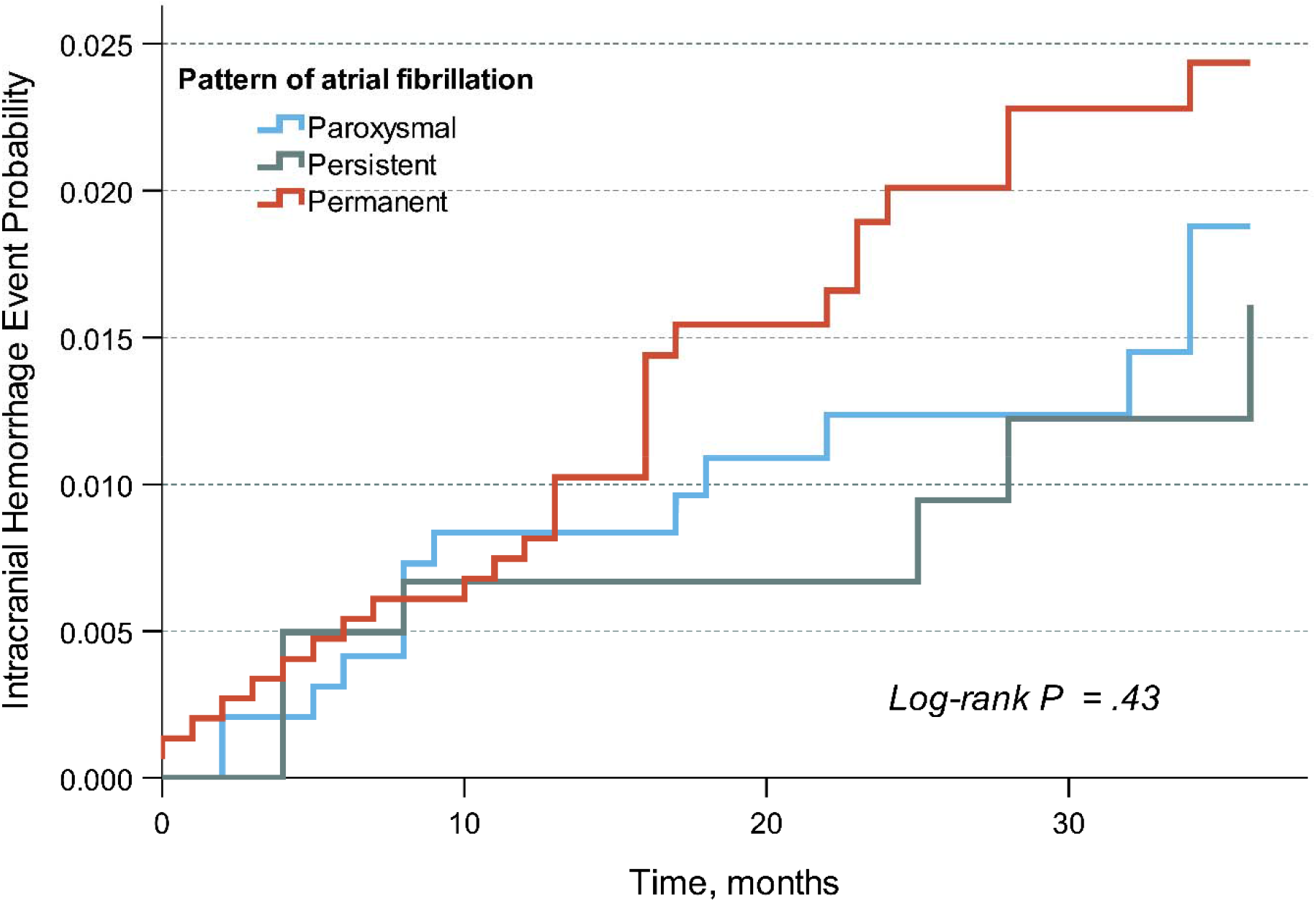
Kaplan-Meier Cumulative Probability Curve of Ischemic Stroke by Pattern of Atrial Fibrillation.

## Discussion

In this large prospective, multicenter cohort of well-anticoagulated AF patients, excess mortality risks were demonstrated in persistent and permanent AF over paroxysmal AF. The increase in mortality risk was independent of age, sex, use of anticoagulation, history of bleeding, and multiple comorbidities. The risks of ischemic stroke, on the other hand, were statistically similar across all patterns of AF.

The prognostic significance of AF pattern has been analyzed in many trials and registries. Among selected, well-controlled, and well-anticoagulated population (ENGAGE AF-TIMI 48,^5^ ROCKET-AF,^20^ and AMADEUS^21^ trials), non-paroxysmal AF was independently associated with worse survival and higher thrombo-embolic event than paroxysmal AF. The results have been conflicting in “real-world” populations. In the Loire valley atrial fibrillation project,^22^ age and comorbidities, rather than pattern of AF, were associated with risk of stroke and all-cause mortality. In the Fushimi^9^ and GARFIELD-AF^23^ registries, the independent association between adverse clinical outcomes and AF pattern were demonstrated but with some dissimilarity. In anticoagulated patients, one registry^9^ showed a prognostic significance of AF pattern in ischemic stroke but not all-cause mortality while a different study showed the contrary.^23^

Besides geographic area, an important difference among these registries and ours was the proportion of non-paroxysmal AF and the anticoagulation rate. We enrolled a higher percentage of non-paroxysmal AF (68%) than the above-mentioned registries (42% in Loire valley registry^22^ and 51% in GARFIELD-AF^23^). The anticoagulation rate in our trial was higher (75% of all patients, 81% of patients with CHA_2_DS_2_-VAS_c_ ≥ 2); compared to 40% in Loire valley registry,^22^ 53% in Fushimi registry,^9^ and 68% in GARFIELD-AF.^23^ We detected no differences in risks of ischemic stroke across all patterns of AF. This finding could likely be explained by the adequacy of anticoagulation in our trial.

There was an approximately 30% increase in mortality risk of non-paroxysmal over paroxysmal AF. Among the non-paroxysmal patterns themselves (persistent and permanent), the risks were statistically similar. Though multiple differences in baseline characteristics were detected between the patterns, they were all adjusted in a well-validated regression model. These result support the concept that non-paroxysmal pattern of AF was not just a term of duration and frequency—rather, it was a disease state—an advanced form associated with worse outcomes.^24^ Early interventions to slow down or prevent progression could be beneficial in newly diagnosed AF. Finally, the increase in mortality risk was not driven by any particular causes of death either cardiovascular or non-cardiovascular. As a matter of fact, non-cardiovascular death occurred more frequently than cardiovascular death suggesting that a more comprehensive approach in AF management is essential to improving outcomes in our patients.

### Strengths

The study design was prospective and multicenter. Data collection was validated and adjudicated. The incidences of the major outcomes were comparable to those reported globally and regionally^9,22,25-27^ reflecting the quality of AF management and the adequacy of event reporting. Finally, unlike most of the clinical trials,^28^ the Cox-regression models in our trial were validated for proportional assumptions.

### Limitations

Due to the nature of the registry, only the association, not the causation, could be demonstrated. Baseline characteristics were not balanced and the therapeutic strategy was not controlled between groups. Despite statistical adjustment, residual confounders remained. Similar to previous trials, AF pattern was defined clinically at physician discretion, which is known to be subject to misclassification error.^5-6,9,22-23^ Vitamin K antagonist was the main choice of anticoagulant. The use of DOAC was limited by the reimbursement policy. Finally, the cause of death was reported as “undetermined” in 20% of all deaths. However, the prognostic significance of any specific causes of death was not the primary research question here.

## Conclusion

In this multicenter cohort of well-anticoagulated AF patients, the temporal pattern of AF was prognostic for all-cause mortality but not for ischemic stroke. Non-paroxysmal AF was associated with an approximately 30% increase in all-cause mortality.

## Data Availability

All data will be provided upon request.

